# Postpartum anxiety and depression symptoms in non-birthing parents in Canada: A cross-sectional study

**DOI:** 10.1101/2025.08.15.25333775

**Authors:** Justine Dol, Christine T. Chambers, Emily Cameron, Cindy-Lee Dennis, Jennifer M. Goldberg, Jennifer A. Parker

## Abstract

**Introduction:** The postpartum period is a vulnerable time for parents. While the focus of most research is on the birthing parent, evidence of postpartum mental health challenges for fathers and sexual minority parents is lacking. The study objective was to determine the prevalence of postpartum depression and anxiety in non-birthing parents, overall and by on sex, gender, and sexual orientation.

**Methods:** An online cross-sectional study was conducted with non-birthing parents from across Canada who had an infant <12 months of age. Recruitment occurred via social media and an online survey company. Parents completed questionnaires, including the Edinburgh Postpartum Depression Scale (EPDS) and General Anxiety Disorder (GAD). Scores >9 and >10 on the EPDS and GAD, respectively, were considered positive for postpartum depression and anxiety symptoms. T-tests were used to determine if there were differences based on sex, gender, or sexual orientation.

**Results:** A total of 133 non-birthing parents participated (54.9% first-time parents, 90.2% fathers). Overall, 56.4% of non-birthing parents had postpartum depression, 23.3% had postpartum anxiety, and 21.8% had comorbid postpartum anxiety and depression. There were no differences based on sex or gender; however, sexual minority parents had a significantly higher prevalence of both postpartum depression (73.3%) and anxiety symptoms (46.7%) than heterosexual parents (52.5% and 16.8%), respectively.

**Discussion:** The postpartum mental health of non-birthing parents is of critical concern with 1 in 2 experiencing symptoms of depression and 1 in 4 experiencing symptoms of anxiety. More work is needed to better support these parents during their first year postpartum.

## INTRODUCTION

Within the postpartum period, much of the clinical care and research is often focused on the transition to parenthood for birthing parents (e.g., cisgendered, heterosexual mothers) [1,2], while evidence on the transition experience for the non-birthing partner (e.g., fathers, sexual minority partners) is lacking [3–5]. The postpartum period, up to 12 months post-birth, is an extremely vulnerable time, with a high risk for parents of developing or experiencing exacerbated mental health issues [6]. Even in uncomplicated pregnancies and births, one in eight mothers and one in ten fathers will experience postpartum mental health concerns [7–10], which can have negative impacts on parenting relationships and child outcomes [11–14].

Non-birthing parents, specifically fathers, often face challenges in their mental health in the postpartum period and frequently feel unsupported and left out by healthcare providers delivering perinatal services [3–5,15–18]. The prevalence of postpartum anxiety and depression in fathers increased during the COVID-19 pandemic, with Canadian estimates around 58.3% for depression and 33.3% for anxiety [19]. However, it is unknown whether this increased prevalence was only during the pandemic or if non-birthing parents continue to experience high levels of mental health struggles after the birth of a child.

There is even less research on perinatal mental health in 2SLGBTQA and sexual minority non-birthing parents [20,21]. This gap in knowledge is despite a growing number of diverse and non-normative family structures, with the percentage of same-sex child-rearing couples in Canada growing from 8.6% in 2001 to 12.0% in 2016 [22]. There is even less research on trans parents, with one study finding that 25% of probability-sampled trans people identified as parents, many experiencing parenting challenges and external stressors [23]. Challenges related to heteronormativity and cisnormativity are consistently identified by sexual minority parents during pregnancy, childbirth, and the postpartum period [6,24,25]. Furthermore, there are differences experienced between sexual minority people who birth children compared to non-birthing parents, with the latter experiencing further exclusion and isolation [6]. 2SLGBTQ+ non-birthing parents can face social exclusion and isolation and lack of support [26], which can contribute to perinatal mental health challenges such as depression and anxiety [27]. Therefore, this study aimed to (1) determine the prevalence of postpartum depression and anxiety in non-birthing parents living in Canada; (2) determine if there is any difference in prevalence based on sex, gender, or sexual orientation.

## METHODS

A cross-sectional online survey was conducted to explore the postpartum experience of non-birthing parents living in Canada. This is part of a larger study exploring the overall postpartum experience for non-birthing parents in Canada.

### Setting & Sample

This study recruited a convenience sample of non-birthing parents. Eligible non-birthing parents included anyone who: (1) identified as a non-birthing parent; (2) had a partner who gave birth within the past 12 months; (3) was over 18 years of age; (4) could read and understand English, and (5) lived in Canada. As this study is exploratory, no sample size calculation was done. Instead, in line with a pragmatic approach that balances resource constraints and feasibility [28,29], we recruited participants over a period of four months to obtain as many responses as possible.

### Data Collection

All data collection occurred remotely via an online survey hosted on Qualtrics with data stored on a secure hospital server. Recruitment occurred through an open call (promoted on social media, posters, and partner outreach) as well as through an online survey company (Leger). Participants who were recruited through the open call between April 3, 2024 and August 14, 2024 had the opportunity to enter a draw for one of five $25 electronic gift cards. Participants were recruited through the survey company between December 4, 2024 and December 10, 2024 and were compensated through the platform.

All participants first completed an online consent and eligibility screening questionnaire before beginning the survey. The survey contained several questionnaires about their postpartum experience as well as demographic information. The survey took approximately 43 minutes to complete (standard deviation (SD): 30.73 minutes, range: 8-196 minutes). Studies completed in full were eligible for inclusion in the analysis. Participants were allowed to stop the survey at any time by exiting; incomplete responses were excluded from the analysis. All data were de-identified prior to analysis to ensure confidentiality.

### Ethical Procedures

Institutional ethical approval was obtained from the IWK Health research ethics board (REB#1029967).

### Outcome Measures

Postpartum depression symptoms were measured using the *Edinburgh Postnatal Depression Scale* (EPDS) [30], a self-report screening scale with 10 items that can indicate if a respondent has symptoms related to perinatal depression [30,31]. The EPDS is valid for assessing depressive symptoms in all people across the perinatal period [32] with higher scores indicating higher symptomology. A score of 9 or higher on the EPDS is the cut-off score indicating probable depression in community-based settings [33]. Postpartum anxiety symptoms were measured using the *Generalized Anxiety Disorder* (GAD-7) scale [34], which includes 7-items to assess generalized anxiety disorder, with scores ranging between 0 and 21. A cut-off score of 10 or greater indicates anxiety at a moderate/severe level. It is important to note that these scales are only screening tools, not diagnostic; however, they were used to reflect measurement tools used in community practice.

### Data Analysis

Quantitative data were analyzed descriptively using Statistical Package for the Social Sciences (SPSS, version 29). Frequencies and other descriptive statistics were used to report demographic characteristics and prevalence rates. Means and standard deviations were calculated to determine the non-birthing parents’ level of postpartum anxiety and depression symptoms. Independent t-tests were used to examine differences between groups based on sex (male, female), gender (cisgender, non-cisgender), or sexual orientation (heterosexual, sexual minority). A p-value of 0.05 was considered statistically significant for all outcomes.

To ensure the validity of the responses to the online survey, best practice strategies were used to mitigate and manage potentially inaccurate responses [35,36]. CAPTCHA, a common security measure for online surveys that helps ensure respondents are human, was used on the consent page to minimize the likelihood of bot attacks. We also set up security checks within the survey itself, including attention checks (i.e., asking questions like *‘If you are reading this, check ‘nearly every day’’* in the middle of a survey), double-checks (i.e., asking the infant’s date of birth twice and participant’s date of birth), and honeypots (i.e., hidden questions that are invisible to humans but automatically completed by bots). Prior to including responses in the analysis, we verified: (1) IP addresses, names, and emails were not duplicated, (2) speed of survey completion to ensure it is realistic (no less than 8 minutes, reflecting 1/3 of estimated completion time); and (3) ensure there were no poor-quality responses (e.g., nonresponse responses to open-ended questions, inconsistent responses across questions). Any responses that had two or more of these concerns were removed from the analysis.

## RESULTS

### Participants

A total of 133 non-birthing parents completed the survey, including 120 (90.2%) who identified as fathers and 13 (9.8%) who identified as non-birthing parents (see Table 1). In terms of sex, 123 identified as male (92.5%); in relation to gender identity, the majority were cisgender men (n=109, 82.0%). Of the 131 parents who identified their sexual orientation, most were heterosexual (n=101, 76.0%).

**Table 1.**
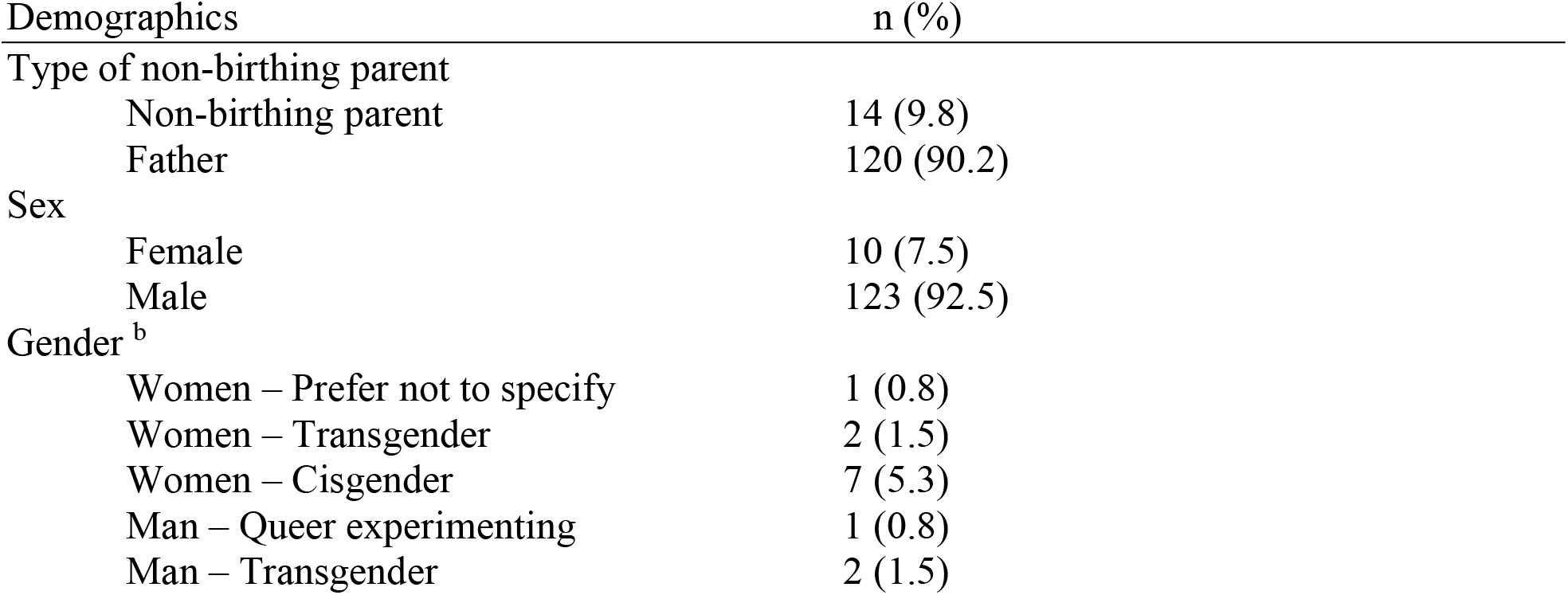
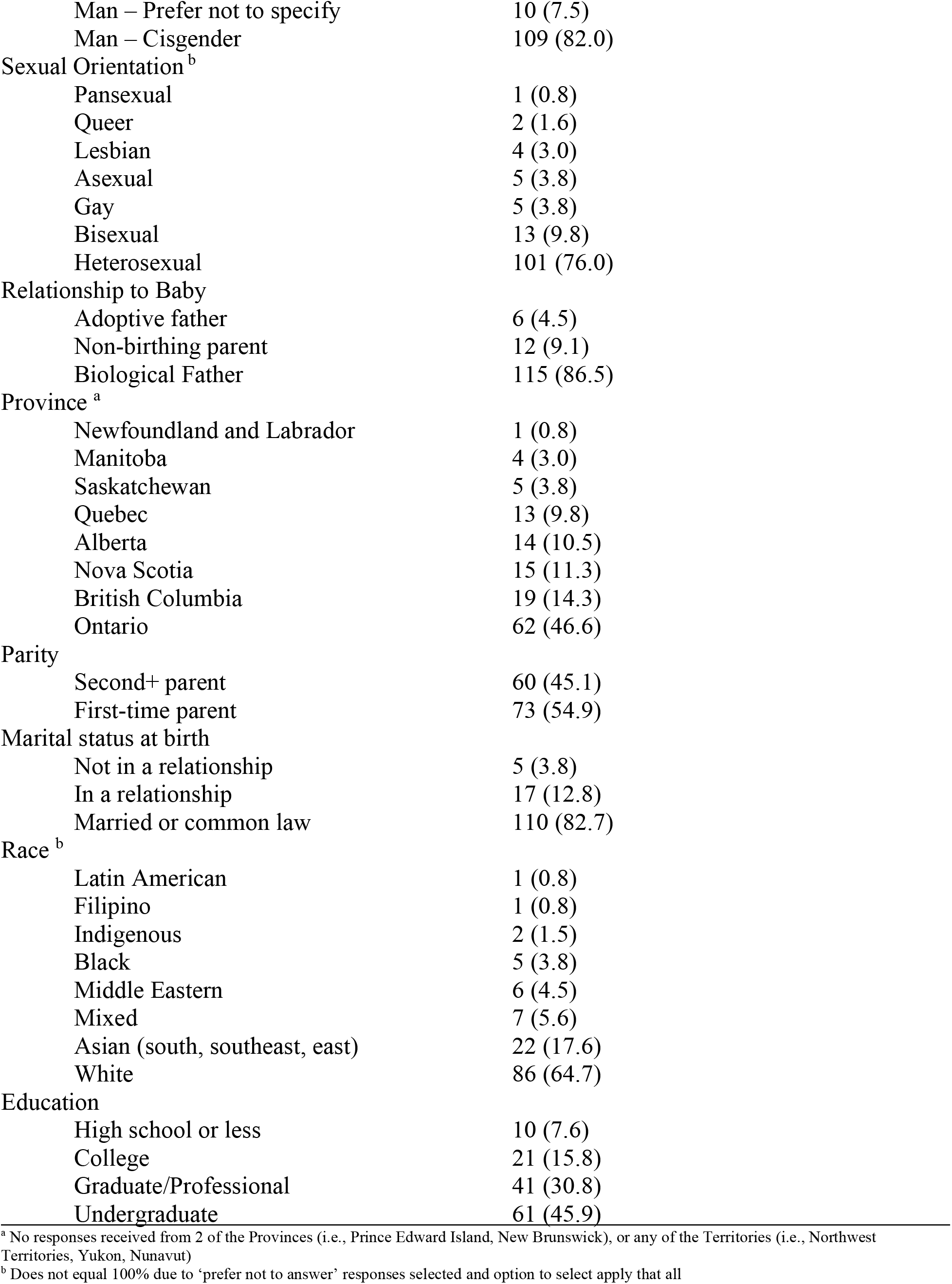
Demographic characteristics of study participants (n=133)

| Demographics | n (%) |
| --- | --- |
| Type of non-birthing parent |  |
| Non-birthing parent | 14 (9.8) |
| Father | 120 (90.2) |
| Sex |  |
| Female | 10 (7.5) |
| Male | 123 (92.5) |
| Gender<sup>b</sup> |  |
| Women – Prefer not to specify | 1 (0.8) |
| Women – Transgender | 2 (1.5) |
| Women – Cisgender | 7 (5.3) |
| Man – Queer experimenting | 1 (0.8) |
| Man – Transgender | 2 (1.5) |

|  |  |  |
| --- | --- | --- |
|  | Man – Prefer not to specify | 10 (7.5) |
|  | Man – Cisgender | 109 (82.0) |
| Sexual Orientation<sup>b</sup> |  |  |
|  | Pansexual | 1 (0.8) |
|  | Queer | 2 (1.6) |
|  | Lesbian | 4 (3.0) |
|  | Asexual | 5 (3.8) |
|  | Gay | 5 (3.8) |
|  | Bisexual | 13 (9.8) |
|  | Heterosexual | 101 (76.0) |
| Relationship to Baby |  |  |
|  | Adoptive father | 6 (4.5) |
|  | Non-birthing parent | 12 (9.1) |
|  | Biological Father | 115 (86.5) |
| Province<sup>a</sup> |  |  |
|  | Newfoundland and Labrador | 1 (0.8) |
|  | Manitoba | 4 (3.0) |
|  | Saskatchewan | 5 (3.8) |
|  | Quebec | 13 (9.8) |
|  | Alberta | 14 (10.5) |
|  | Nova Scotia | 15 (11.3) |
|  | British Columbia | 19 (14.3) |
|  | Ontario | 62 (46.6) |
| Parity |  |  |
|  | Second+ parent | 60 (45.1) |
|  | First-time parent | 73 (54.9) |
| Marital status at birth |  |  |
|  | Not in a relationship | 5 (3.8) |
|  | In a relationship | 17 (12.8) |
|  | Married or common law | 110 (82.7) |
| Race<sup>b</sup> |  |  |
|  | Latin American | 1 (0.8) |
|  | Filipino | 1 (0.8) |
|  | Indigenous | 2 (1.5) |
|  | Black | 5 (3.8) |
|  | Middle Eastern | 6 (4.5) |
|  | Mixed | 7 (5.6) |
|  | Asian (south, southeast, east) | 22 (17.6) |
|  | White | 86 (64.7) |
| Education |  |  |
|  | High school or less | 10 (7.6) |
|  | College | 21 (15.8) |
|  | Graduate/Professional | 41 (30.8) |
|  | Undergraduate | 61 (45.9) |

<sup>a</sup> No responses received from 2 of the Provinces (i.e., Prince Edward Island, New Brunswick), or any of the Territories (i.e., Northwest Territories, Yukon, Nunavut)
<sup>b</sup> Does not equal 100% due to 'prefer not to answer' responses selected and option to select apply that all

Participants reported a mean age of 35.6 years (SD=6.0 years) and their children were on average 6.5 months old at the time of survey completion (SD=3.5 months, range: 0-12 months). Overall, non-birthing parents, most identified as the biological father (n=115, 86.5%) and 54.9% were experiencing parenthood for the first time. Non-birthing parents were predominantly white (n=86, 64.7%), had an undergraduate degree (n=61, 45.9%), and were from Ontario (n=62, 46.6%), followed by British Columbia (n=19, 14.3%).

Across all non-birthing parents, 56.4% (n=75) met the clinical cut-off for postpartum depression symptoms (M=9.5, SD=5.9) and 23.3% (n=31) for postpartum anxiety symptoms (M=5.8, SD=5.3, Table 2). While 42.1% of non-birthing parents did not have either postpartum depression or anxiety symptoms, 21.8% of non-birthing parents had clinical levels of comorbid postpartum anxiety and depression symptoms. These prevalence rates remained consistent when considering geographical regions within Canada (Supplementary Table S1 and S2).

**Table 2.** Prevalence of postpartum depression and anxiety symptoms in non-birthing parents.

| **Outcome** | **Non-birthing**<br>**parents (N=133)**<br>**N (%)** | **Sexual minority**<br>**parent (n=30)**<br>**N (%)** | **Heterosexual**<br>**parent (n=101)**<br>**N (%)** | **p value** |
| --- | --- | --- | --- | --- |
| Postpartum anxiety | 31 (23.3) | 14 (46.7) | 17 (16.8) | < 0.001 |
| Postpartum depression | 75 (56.4) | 22 (73.3) | 53 (52.5) | 0.043 |

When examining by sex and gender, there were no differences between groups on postpartum anxiety or depression symptoms or prevalence rates (Supplementary Tables S3 and S4). However, non-birthing parents who self-identified as a sexual minority parent (Table 1) had significantly higher postpartum depression and anxiety symptoms than those who identified as heterosexual (Table 3). Furthermore, as shown in Table 2, sexual minority non-birthing parents have a significantly higher prevalence of both postpartum depression (73.3%) and anxiety symptoms (46.7%) than heterosexual parents (52.5% and 16.8%, respectively).

**Table 3.** Postpartum depression and anxiety symptom scores based on sexual orientation of non-birthing parent.

| **Outcome** | **Sexual Minority (n=30)**<br>**Mean (SD)** | **Heterosexual (n=101)**<br>**Mean (SD)** | **p value** |
| --- | --- | --- | --- |
| Postpartum anxiety symptoms | 8.80 (5.93) | 5.12 (4.74) | 0.01 |
| Postpartum depression symptoms | 12.23 (6.09) | 8.88 (5.68) | 0.003 |

## DISCUSSION

This cross-sectional study sought to determine the prevalence of postpartum anxiety and depression symptoms in non-birthing parents across Canada while also determining if there were any differences in prevalence based on sex, gender, and sexual orientation. Overall, over half of non-birthing parents surveyed had postpartum depression symptoms, almost a quarter had postpartum anxiety symptoms, and one-fifth had comorbid postpartum anxiety and depression symptoms. While there were no differences based on sex or gender, parents who identified as a sexual minority parent (e.g., bisexual, asexual, lesbian, gay, queer, or pansexual) had significantly higher postpartum anxiety and depression symptoms than those who identified as heterosexual. This research highlights not only the high prevalence of postpartum depression and anxiety symptoms in non-birthing parents across the first year postpartum broadly, but also for sexual minority parents specifically.

Previous meta-analytic work found that between 5.6% to 8.4% of fathers experience postpartum depression and 9.5% of fathers experience postpartum anxiety [37,38]. In a recent Canadian study, 22.4% of fathers had comorbid anxiety and depression at some time during the first year postpartum, with approximately 11% experiencing postpartum depression and 22% experiencing postpartum anxiety symptoms between 3-12 months [39]. In the current study, postpartum depression prevalence was considerably higher than these findings, even when separated by sex, gender, and sexual orientation. Compared to Dennis et al. [39], our study found similar rates of postpartum anxiety across approximately a quarter of all non-birthing parents. This suggests that non-birthing parents are experiencing high rates of postpartum anxiety and depression symptoms which may not be managed effectively. Interestingly, data from these studies anxiety [37–39] were collected prior to the COVID-19 pandemic, with recent work suggesting that fathers experienced higher rates of postpartum anxiety and depression during the pandemic than before [40]. In a study conducted early in the pandemic in 2020, 58.3% of Canadian fathers with a child between the ages of 0 and 18 months had postpartum depression and 33.3% had postpartum anxiety [19]. These figures are more consistent with our findings, suggesting that postpartum anxiety and depression prevalence rates remain high post-pandemic and there is a need to better integrate postpartum mental health support for non-birthing parents.

Our study builds on the limited evidence available on the prevalence of postpartum anxiety and depression for sexual minority non-birthing parents. Our findings indicate that sexual minority non-birthing parents are experiencing significantly higher rates of both postpartum anxiety and depression compared to heterosexual fathers. Previous work has found that sexual minority women (e.g., lesbian, bisexual) had higher postpartum depression scores compared to heterosexual women [41–46]. Postpartum depression for gay fathers is approximately 12% [47], with evidence suggesting that the experiences of gay fathers is distinct from heterosexual fathers yet infrequently studied [48,49]. This suggests that there may be a multitude of factors within the sexual minority non-birthing parent population that influence postpartum mental health. Further research is needed to understand why sexual minority non-birthing parents not only experience higher postpartum mental health challenges but also to examine differences within this heterogeneous group. Other reviews have identified similar gaps in the literature, highlighting the lack of evidence on perinatal mental health in transgender and non-binary parents (Greenfield and Darwin, 2020) and limited evidence on how race, ethnicity, and sexuality intersect to impact non-birthing parents’ mental health [51].

### Clinical Implications

Given the high rate of postpartum anxiety and depression symptoms among non-birthing parents in their first year after childbirth, it is essential to improve mental health screening and support specifically for this often-overlooked group. Addressing this need is vital not only for the well-being of non-birthing parents—by offering timely, appropriate interventions, treatments, and ongoing support—but also for maintaining a healthy family dynamic. Mental health struggles in one parent can create additional stressors and obstacles that hinder the birthing parent’s recovery and impact the child’s emotional and developmental progress. Research confirms that co-parenting maternal and partner mental health are linked, and untreated issues in one caregiver can adversely affect parenting, the couple’s relationship, and the child’s overall health [51]. There is a need to focus on postpartum mental health not as a single parent issue, but one that impacts both parents and therefore the family unit.

To effectively address this issue, it is essential to enhance education and awareness efforts at multiple levels. Healthcare providers require specific training and tools to help them identify and screen for postpartum mental health issues in non-birthing parents. Besides individual screenings, educational initiatives should aim to normalize postpartum mental health challenges for all types of parents, including fathers, partners, and those outside traditional heterosexual or cisgender roles, so that non-birthing parents feel more comfortable discussing their experiences without fear of stigma or dismissal. Additionally, specialized training is particularly important for healthcare teams to reduce heteronormative assumptions and unconscious biases that could lead to structural discrimination against sexual minority parents [52]. By actively fostering more inclusive and affirming healthcare environments, we can help ensure that all parents receive the mental health support they need during the postpartum period, ultimately supporting healthier and more resilient family systems.

### Strengths and Limitations

The primary strength of this study lies in its inclusive approach to identifying non-birthing parents, which fills a gap in the literature. Including non-birthing parents across the first 12 months postpartum anywhere in Canada provides a comprehensive picture of postpartum anxiety and depression symptoms in non-birthing parents across sexual identities. However, several limitations should be acknowledged. First, despite attempts to recruit non-birthing parents to complete the survey, we were only able to collect 133 eligible responses during the period of data collection. This challenge has been identified by others researching non-birthing parents in the postpartum period [19], and opportunities to expand targeted data collection is warranted. Furthermore, within this sample, most non-birthing parents identified as male, high socioeconomic status, cisgender, and heterosexual. There was also a lack of diversity in racial identities. Our findings should be interpreted with this in mind as results may not be generalizable to groups who experience multiple intersecting oppressions. More research is needed on non-birthing parents outside this limited demographic to ensure that their voices are heard, and their experiences are understood. Another limitation is that most participants were from Ontario, with no representation from Prince Edward Island, New Brunswick, or the Territories, nor was information on rural versus urban collected, limiting the representation and deeper analysis based on geographical location. Finally, our sample of adoptive parents was extremely small, yet evidence suggests that important differences exist for parents who adopt compared to biological parents, which warrants further attention [53].

## Conclusions

In this cross-sectional study, non-birthing parents reported high prevalence rates of both postpartum anxiety and depression, with sexual minority non-birthing parents experiencing higher rates than heterosexual non-birthing parents. Postpartum mental health of non-birthing parents is of critical concern, with more research needed to understand how to support non-birthing parents during the first year postpartum and beyond.

## Data Availability

The data that support the findings of this study are available from the corresponding author, JD, upon reasonable request.

## Author Contribution

**Conceptualization:** JD, CTC, JAP, CLD

**Data Curation:** JD

**Funding Acquisition:** JD, CTC,

**Methodology:** JD, CTC, JAP, CLD

**Project Administration:** JD

**Resources**: CTC

**Supervision**: CTC, JAP

**Visualization**: JD

**Writing – Original Draft Preparation**: JD

**Writing – Review & Editing:** All

## SUPPLEMENTAL MATERIAL

**Table S1.** Postpartum depression symptomatology prevalence by Canadian region.

| **Outcome** | **Non-birthing**<br>**parents**<br>**N (%)** | **Sexual minority**<br>**parent n/N (%)** | **Heterosexual parent**<br>**n/N (%)** |
| --- | --- | --- | --- |
| Atlantic Provinces <sup>a</sup><br>(n=16) | 6 (37.5) | 2/2 (100) | 4/14 (28.6) |
| Central Canada <sup>b</sup><br>(n=75) | 44 (58.7) | 12/18 (66.7) | 32/56 (57.1) |
| Prairie Provinces <sup>c</sup><br>(n=23) | 15 (65.2) | 5/7 (71.4) | 10/15 (66.7) |
| West Coast <sup>d</sup><br>(n=19) | 10 (52.6) | 3/3 (100) | 7/16 (43.8) |

<sup>a</sup> Newfoundland and Labrador, Prince Edward Island, Nova Scotia, and New Brunswick
<sup>b</sup> Ontario and Quebec
<sup>c</sup> Manitoba, Saskatchewan, and Alberta
<sup>d</sup> British Columbia
N.B. Northern Territories not reported on due to lack of respondents

**Table S2.** Postpartum anxiety symptomatology prevalence by Canadian region.

| **Outcome** | **Non-birthing**<br>**parents**<br>**N (%)** | **Sexual minority**<br>**parent n (%)** | **Heterosexual parent**<br>**n (%)** |
| --- | --- | --- | --- |
| Atlantic Provinces <sup>a</sup><br>(n=16) | 3 (18.8) | 1/2 (50.0) | 2/14 (14.3) |
| Central Canada <sup>b</sup><br>(n=75) | 19 (25.3) | 9/18 (50.0) | 10/56 (17.9) |
| Prairie Provinces <sup>c</sup><br>(n=23) | 4 (17.3) | 1/7 (14.3) | 3/15 (20.0) |
| West Coast <sup>d</sup><br>(n=19) | 5 (26.3) | 3/3 (100) | 2/16 (12.5) |

<sup>a</sup> Newfoundland and Labrador, Prince Edward Island, Nova Scotia, and New Brunswick
<sup>b</sup> Ontario and Quebec
<sup>c</sup> Manitoba, Saskatchewan, and Alberta
<sup>d</sup> British Columbia
N.B. Northern Territories not reported on due to lack of respondents

**Table S3.** Postpartum depression and anxiety scores and prevalence based on sex.

| **Outcome** | **Female (n=10)** |  | **Male (n=123)** |  |
| --- | --- | --- | --- | --- |
|  | **Score**<br>**Mean (SD)** | **Prevalence**<br>**N (%)** | **Score**<br>**Mean (SD)** | **Prevalence**<br>**N (%)** |
| Postpartum anxiety | 7.40 (6.63) | 4 (40.0) | 5.75 (5.14) | 27 (22.0) |
| Postpartum depression | 9.60 (6.11) | 6 (60.0) | 9.54 (5.96) | 69 (56.1) |

**Table S4.** Postpartum depression and anxiety scores and prevalence based on gender.

| **Outcome** | **Non-cisgendered (n=16)** |  | **Cisgender (n=116)** |  |
| --- | --- | --- | --- | --- |
|  | **Score**<br>**Mean (SD)** | **Prevalence**<br>**N (%)** | **Score**<br>**Mean (SD)** | **Prevalence**<br>**N (%)** |
| Postpartum anxiety | 6.06 (5.95) | 5 (31.3) | 5.90 (5.18) | 26 (22.4) |
| Postpartum depression | 9.19 (7.10) | 8 (50.0) | 9.66 (5.78) | 67 (57.8) |

## References

[1] Prinds C, Hvidt NC, Mogensen O, Buus N. Making existential meaning in transition to motherhood-A scoping review. Midwifery 2014;30:733–41. 10.1016/j.midw.2013.06.021.

[2] Walker SB, Rossi DM, Sander TM. Women’s successful transition to motherhood during the early postnatal period: A qualitative systematic review of postnatal and midwifery home care literature. Midwifery 2019;79:102552. 10.1016/j.midw.2019.102552.

[3] Kowlessar O, Fox JR, Wittkowski A. First-time fathers’ experiences of parenting during the first year. J Reprod Infant Psychol 2015;33:4–14. 10.1080/02646838.2014.971404.

[4] Chin R, Daiches A, Hall P. A qualitative exploration of first-time fathers’ experiences of becoming a father. Community Pract J Community Pract Health Visit Assoc 2011;84:19–23.

[5] McKelvey MM. The other mother: A narrative analysis of the postpartum experiences of nonbirth lesbian mothers. Adv Nurs Sci 2014;37:101–16. 10.1097/ANS.0000000000000022.

[6] Miller ML, Dupree J, Monette MA, Lau EK, Peipert A. Health Equity and Perinatal Mental Health. Curr Psychiatry Rep 2024;26:460–9. 10.1007/s11920-024-01521-4.

[7] Leach LS, Poyser C, Cooklin AR, Giallo R. Prevalence and course of anxiety disorders (and symptom levels) in men across the perinatal period: A systematic review. J Affect Disord 2016;190:675–86. 10.1016/j.jad.2015.09.063.

[8] Paulson J, Bazemore S. Prenatal and postpartum depression in fathers and its association with maternal depression: A meta-analysis. JAMA 2010;303:1961–9.

[9] Hetherington E, McDonald S, Williamson T, Patten SB, Tough SC. Social support and maternal mental health at 4 months and 1 year postpartum: analysis from the All Our Families cohort. J Epidemiol Community Health 2018;72:933–9. 10.1136/jech-2017-210274.

[10] Dennis CLL, Falah-Hassani K, Brown HK, Vigod SN. Identifying women at risk for postpartum anxiety: a prospective population-based study. Acta Psychiatr Scand 2016;134:485–93. 10.1111/acps.12648.

[11] Dachew B, Ayano G, Duko B, Lawrence B, Betts K, Alati R. Paternal Depression and Risk of Depression Among Offspring: A Systematic Review and Meta-Analysis. JAMA Netw Open 2023;6:e2329159. 10.1001/jamanetworkopen.2023.29159.

[12] Ross L. Perinatal Mental Health in Lesbian Mothers: A Review of Potential Risk and Protective Factors. Women Health 2005;41:113–28. 10.1300/J013v41n03.

[13] Sweeney S, MacBeth A. The effects of paternal depression on child and adolescent outcomes: A systematic review. J Affect Disord 2016;205:44–59. 10.1016/j.jad.2016.05.073.

[14] Wilson S, Durbin CE. Effects of paternal depression on fathers’ parenting behaviors: a meta-analytic review. Clin Psychol Rev 2010;30:167–80. 10.1016/j.cpr.2009.10.007.

[15] Pollock MA, Amankwaa LC, Amankwaa AA. First-Time Fathers and Stressors in the Postpartum Period. J Perinat Educ 2005;14:19–25. 10.1624/105812405×44682.

[16] Hudson DB, Elek SM, Fleck MOFE. First-time mothers’ and fathers’ transition to parenthood: Infant care, self-efficacy, parenting satisfcation, and infant sex. Compr Pediatr Nurs 2001;24:31–43.

[17] StGeorge JM, Fletcher RJ, St George JM, Fletcher RJ, StGeorge JM, Fletcher RJ. Fathers Online: Learning About Fatherhood Through the Internet. J Perinat Educ 2011;20:154–62. 10.1891/1058-1243.20.3.154.

[18] Åsenhed L, Kilstam J, Alehagen S, Baggens C. Becoming a father is an emotional roller coaster -an analysis of first-time fathers′ blogs. J Clin Nurs 2014;23:1309–17. 10.1111/jocn.12355.

[19] Cameron EE, Joyce KM, Hatherly K, Roos LE. Paternal Depression and Anxiety During the COVID-19 Pandemic. Int J Environ Res Public Health 2025;22:124. 10.3390/ijerph22010124.

[20] Hafford-Letchfield T, Cocker C, Rutter D, Tinarwo M, McCormack K, Manning R. What do we know about transgender parenting?: Findings from a systematic review. Health Soc Care Community 2019;27:1111–25. 10.1111/hsc.12759.

[21] Biocco R, Carone N, Ioverno S, Lingiardi V. Same-Sex and Different-Sex Parent Families in Italy: Is Parents’ Sexual Orientation Associated with Child Health Outcomes and Parental Dimensions? J Dev Behav Pediatr JDBP 2018;39:555–63.

[22] Statistics Canada. Census in Brief: Same-sex couples in Canada in 2016. Stat Can 2017. https://www12.statcan.gc.ca/census-recensement/2016/as-sa/98-200-x/2016007/98-200-x2016007-eng.cfm (accessed July 23, 2020).

[23] Pyne J, Bauer G, Bradley K. Transphobia and Other Stressors Impacting Trans Parents. J GLBT Fam Stud 2015;11:107–26. 10.1080/1550428X.2014.941127.

[24] Goldberg JM, Ross LE. Attitudes of Midwives Towards Lesbians: Results From a Systematic Review of Literature on Midwives’ Attitudes Towards Sexual and Gender Minority People. Can J Midwifery Res Pract 2022;21. 10.22374/cjmrp.v21i1.1.

[25] Stewart K, O’Reilly P. Exploring the attitudes, knowledge and beliefs of nurses and midwives of the healthcare needs of the LGBTQ population: An integrative review. Nurse Educ Today 2017;53:67–77. 10.1016/j.nedt.2017.04.008.

[26] Hudak N. “Who’s the Mom?”: Heterosexism in Patient-Provider Interactions of Queer Pregnant Couples. Health Commun 2023;38:114–23. 10.1080/10410236.2021.1936752.

[27] Howat A, Masterson C, Darwin Z. Non-birthing mothers’ experiences of perinatal anxiety and depression: Understanding the perspectives of the non-birthing mothers in female same-sex parented families. Midwifery 2023;120:103650. 10.1016/j.midw.2023.103650.

[28] Foster C. Methodological pragmatism in educational research: from qualitative-quantitative to exploratory-confirmatory distinctions. Int J Res Method Educ 2024;47:4–19. 10.1080/1743727X.2023.2210063.

[29] Yaremych HE, Persky S. Recruiting Fathers for Parenting Research: An Evaluation of Eight Recruitment Methods and an Exploration of Fathers’ Motivations for Participation. Parent Sci Pract 2023;23:1–32. 10.1080/15295192.2022.2036940.

[30] Cox J, Holden J, Sagovksy R. Detection of Postnatal Depression: Development of the 10-item Edinburgh Postnatal Depression Scale. Br J Psychiatry 1987;150:782–6. 10.1007/978-94-007-1694-0_2.

[31] Cox J, Holden J, Henshaw C. The Edinburgh Post Natal Depression Scale (EPDS) Manual. 2014.

[32] Edmondson OJH, Psychogiou L, Vlachos H, Netsi E, Ramchandani PG. Depression in fathers in the postnatal period: Assessment of the Edinburgh Postnatal Depression Scale as a screening measure. J Affect Disord 2010;125:365–8. 10.1016/j.jad.2010.01.069.

[33] Matthey S, Henshaw C, Elliott S, Barnett B. Variability in use of cut-off scores and formats on the Edinburgh Postnatal Depression Scale: implications for clinical and research practice. Arch Womens Ment Health 2006;9:309–15. 10.1007/s00737-006-0152-x.

[34] Spitzer RL, Kroenke K, Williams JBW, Löwe B. A brief measure for assessing generalized anxiety disorder: The GAD-7. Arch Intern Med 2006;166:1092–7. 10.1001/archinte.166.10.1092.

[35] Storozuk A, Ashley M, Delage V, Maloney EA. Got Bots? Practical Recommendations to Protect Online Survey Data from Bot Attacks. Quant Methods Psychol 2020;16:472–81. 10.20982/tqmp.16.5.p472.

[36] Yarrish C, Groshon L, Mitchell JD, Appelbaum A, Klock S, Winternitz T, et al. Finding the signal in the noise: Minimizing responses from bots and inattentive humans in online research. Behav Ther 2019;42:235– 42.

[37] Cameron EE, Sedov ID, Tomfohr-Madsen LM. Prevalence of paternal depression in pregnancy and the postpartum: An updated meta-analysis. J Affect Disord 2016;206:189–203. 10.1016/j.jad.2016.07.044.

[38] Leiferman JA, Farewell CV, Jewell J, Lacy R, Walls J, Harnke B, et al. Anxiety among fathers during the prenatal and postpartum period: a meta-analysis. J Psychosom Obstet Gynecol 2021;0:1–10. 10.1080/0167482X.2021.1885025.

[39] Dennis CL, Marini F, Dol J, Vigod SN, Grigoriadis S, Brown HK. Paternal prevalence and risk factors for comorbid depression and anxiety across the first 2 years postpartum: A nationwide Canadian cohort study. Depress Anxiety 2021. 10.1002/da.23234.

[40] Xavier JF, Perumal RV, Poornaselvan C, Adil M. Perinatal Fathers in the Context of the COVID-19 Pandemic and Beyond: Impacts and Implications. Malays J Nurs MJN 2024;16:230–43. 10.31674/mjn.2024.v16i02.023.

[41] Flanders CE, Gibson MF, Goldberg AE, Ross LE. Postpartum depression among visible and invisible sexual minority women: a pilot study. Arch Womens Ment Health 2016;19:299–305. 10.1007/s00737-015-0566-4.

[42] Kirubarajan A, Barker LC, Leung S, Ross LE, Zaheer J, Park B, et al. LGBTQ2S+ childbearing individuals and perinatal mental health: A systematic review. BJOG Int J Obstet Gynaecol 2022;129:1630–43. 10.1111/1471-0528.17103.

[43] Mamrath S, Greenfield M, Fernandez Turienzo C, Fallon V, Silverio SA. Experiences of postpartum anxiety during the COVID-19 pandemic: A mixed methods study and demographic analysis. PLoS One 2024;19:e0297454.

[44] Ross L, Goldberg AE. Perinatal experiences of lesbian, gay, bisexual, and transgender people. Oxf. Handb. Perinat. Psychol., New York, NY, US: Oxford University Press; 2016, p. 618–30.

[45] Ross LE, Steele L, Goldfinger C, Strike C. Perinatal depressive symptomatology among lesbian and bisexual women. Arch Womens Ment Health 2007;10:53–9. 10.1007/s00737-007-0168-x.

[46] Tam M, Goldberg J, Andrade-Romo Z, Ross L. 2SLGBTQ+ Individuals and Perinatal Mental Health Disorders. Routledge Int. Handb. Perinat. Ment. Health Disord., 2024, p. 694–672.

[47] Adler L, Yehoshua I, Mizrahi Reuveni M. Postpartum Depression Among Gay Fathers With Children Born Through Surrogacy: A Cross-sectional Study. J Psychiatr Pract 2023;29:3. 10.1097/PRA.0000000000000684.

[48] Carneiro FA, Tasker F, Salinas-Quiroz F, Leal I, Costa PA. Are the Fathers Alright? A Systematic and Critical Review of Studies on Gay and Bisexual Fatherhood. Front Psychol 2017;8. 10.3389/fpsyg.2017.01636.

[49] Shenkman G, Levy S, Winkler ZB-D, Bass D, Geller S. Higher Levels of Postnatal Depressive Symptomatology, Post-Traumatic Growth, and Life Satisfaction among Gay Fathers through Surrogacy in Comparison to Heterosexual Fathers: A Study in Israel in Times of COVID-19. Int J Environ Res Public Health 2022;19:7946. 10.3390/ijerph19137946.

[50] Greenfield M, and Darwin Z. Trans and non-binary pregnancy, traumatic birth, and perinatal mental health: a scoping review. Int J Transgender Health 2020;22:203–16. 10.1080/26895269.2020.1841057.

[51] Fisher SD, Walsh T, Wongwai C. The importance of perinatal non-birthing parents’ mental health and involvement for family health. Semin Perinatol 2024;48:151950. 10.1016/j.semperi.2024.151950.

[52] Adams C, Bennetts S, Ridgway L, Hooker L, East C, Edvardsson K. Father and non-birth parent experience of child and family health services: a systematic review and meta-synthesis. Aust J Prim Health 2025;31. 10.1071/PY24228.

[53] Long T, Jones C, Jomeen J, Martin CR. Becoming parents by adoption: A systematic review. J Health Visit 2021;9:116–27. 10.12968/johv.2021.9.3.116.

